# Validation of the Croatian Versions of DASH, PRWE and Mayo Wrist Score in patients with Distal Radius Fractures

**DOI:** 10.1101/2025.10.02.25336703

**Authors:** Borjan Josifovski, Matej Pedisic, Mirela Vuckovic, Ksenija Bazdaric, Zdravko Jotanovic

## Abstract

**Introduction:** Distal radius fractures are common upper extremity injuries requiring reliable outcome measures for accurate clinical assessment. This study aimed to validate the Croatian versions of the Disabilities of the Arm, Shoulder and Hand (DASH), Patient-Rated Wrist Evaluation (PRWE) and Mayo Wrist Score (MWS) in patients with distal radius fractures.

**Methods:** DASH, PRWE and MWS croatian versions validity and reliability were evaluated in 128 patients using standardized translation, cultural adaptation, factor analysis, and internal consistency (Cronbach’s α).

**Results:** PRWE-Cro and DASH-Cro demonstrated excellent validity and internal consistency (α > 0.95). QuickDASH-Cro showed high reliability and is recommended as a practical alternative. MWS was validated for the first time, showing good validity and acceptable internal consistency (α = 0.71).

**Conclusion:** PRWE-Cro and DASH-Cro are validated, reliable instruments suitable for both clinical practice and research in Croatian-speaking populations while MWS can be used as a quick screening tool.

## INTRODUCTION

Distal radius fractures (DRFs) are among the most common fractures, accounting for 17.5% of all adult fractures (Court-Brown and Caesar, 2006). Functional outcomes following treatment of these injuries can vary widely, and objective clinical assessments - such as range of motion, grip strength, and radiographic measurements - often fail to capture the patient’s perspective on pain, disability, and return to function. This highlights the need for validated questionnaires that comprehensively integrate clinical examination findings with assessments of activities of daily living. In routine clinical practice, it is essential that the selected questionnaire be simple and understandable for patients, time-efficient, reliable, and suitable for use in scientific research. Consequently, the use of patient-reported outcome measures (PROMs) has become essential in both clinical practice and research for evaluating treatment effectiveness and guiding patient-centered care (Black, 2013). PROMs provide valuable insights into patients’ perceptions of symptoms, functional limitations, and overall quality of life (Black, 2013).

Over the past two decades, PROMs for the functional assessment of the upper extremity have been increasingly developed, aiming to objectify the patient’s clinical status and enable functional monitoring over time (Gabel, 2006). For hand and wrist conditions, a total of 22 specific PROMs have been identified in the literature (Iskander et al., 2025). Among these, the most commonly used are the Disabilities of the Arm, Shoulder and Hand (DASH) questionnaire, the Patient-Rated Wrist (and Hand) Evaluation [PRW(H)E], and the QuickDASH (Iskander et al., 2025). These PROMs are most frequently applied in studies focusing on traumatic hand and wrist conditions (Iskander et al., 2025). A recent study demonstrated that using the QuickDASH and PRWHE alongside the Mayo Wrist Score (MWS) is a critical approach for assessing pain and functional outcomes in patients with distal radius fractures (Ille et al., 2025).

The DASH is a region-specific instrument consisting of 30 items that assess disability and symptoms in the upper extremity across a wide range of conditions, and it is among the most frequently used outcome tools in orthopaedic research and practice (Hudak et al., 1996; Aman et al., 2025). The QuickDASH is a shortened version of the DASH (11 items) designed to measure physical function and symptoms in patients with any musculoskeletal disorder of the upper limb (Beaton et al., 2005; Hammer et al., 2022). The PRWE is a joint-specific instrument with 15 items focused on wrist pain and function, and it has demonstrated excellent responsiveness in patients with distal radius fractures (MacDermid, 1996). The MWS is a brief, 4-item instrument incorporating both clinician and patient input, and it is widely used as a PROM in wrist trauma studies due to its ease of application in clinical settings (Cooney et al., 1987; Dacombe et al., 2016). Nevertheless, to the best of our knowledge, MWS has not been formally validated. Existing literature critiques its mixed subjective-objective format and highlights the lack of established reliability and validity compared to other PROMs such as the PRWE and DASH (Dacombe et al., 2016).

Despite their widespread use and proven utility in English-speaking populations, the applicability of PROMs across different languages and cultural settings requires systematic translation and validation procedures (Tsang et al., 2017). Without cultural adaptation, direct translations may compromise content validity and result in inaccurate assessments. The lack of validated Croatian versions of the DASH, PRWE, and MWS has limited their routine use in clinical practice and research among Croatian-speaking populations.

This study aimed to translate and culturally adapt the DASH, PRWE, and MWS into the Croatian language, and to evaluate their psychometric properties - specifically validity and reliability - in patients with distal radius fractures.

## METHODS

### Study design

A cross-sectional clinimetric study was conducted from January 30, 2024, to February 28, 2025, at the University Hospital for Orthopaedics and Traumatology in Lovran, Croatia, and the Kantrida Nursing Home in Rijeka, Croatia. Ethics approval was obtained from the University Hospital for Orthopaedics and Traumatology in Lovran, Croatia, and the Faculty of Medicine at the University of Rijeka, Croatia, and authorization was obtained from the Kantrida Nursing Home in Rijeka, Croatia. Written informed consent was obtained from all participants prior to study commencement. All collected data were anonymized and stored in a secure database for subsequent statistical analysis.

### Study participants

A total of 128 patients with DRFs completed PROMs. Both conservatively and surgically treated patients were included.

### Inclusion and Exclusion Criteria

The study included patients aged 18 to 90 years, regardless of gender, who had sustained a distal radius fracture at least three months prior to enrollment. Exclusion criteria were: age under 18 or over 90 years, incomplete questionnaire responses, less than three months between the fracture and study enrollment, cognitive impairment, legal guardianship, or insufficient understanding of the Croatian language.

### Procedure

The first group of participants (n = 115) was enrolled during follow-up visits at the University Hospital for Orthopaedics and Traumatology in Lovran, Croatia. The second group (n = 13) consisted of residents of the Kantrida Nursing Home for the Elderly in Rijeka, Croatia, who had sustained distal radius fractures and met the study criteria. These individuals were approached directly at the care facility with the cooperation of healthcare staff. Only participants from both groups who met the inclusion criteria were enrolled in the study. All participants received a detailed explanation of the study’s purpose, and written informed consent was obtained. They were then invited to complete the questionnaires, with authors BJ and MP present to provide clarification and ensure completeness. This supervised administration helped reduce misunderstandings and minimize missing data.

### Translation and adaptation

The cross-cultural adaptation of the PRWE, DASH, and MWS questionnaires was performed according to established guidelines for translation and validation (Tsang et al., 2017). The process began with the forward translation of the original questionnaires into Croatian, independently carried out by two bilingual translators whose native language is Croatian. Discrepancies between the translations were resolved through discussion and consensus in collaboration with two orthopaedic surgeons.

The agreed-upon Croatian versions were then back-translated into English by two independent native English speakers, both blinded to the original versions. A review panel - comprising one of the initial translators, one of the back-translators, a methodology expert in outcome measures, a physiotherapist, and two orthopaedic surgeons (one specializing in hand and wrist surgery) - evaluated all versions. Any inconsistencies identified during the review were resolved by consensus.

A pre-final version of each questionnaire was then developed and pilot-tested on a sample of five patients with distal radius fractures. Based on their feedback, minor adjustments were made, resulting in the final Croatian versions of the PRWE (PRWE-Cro), DASH (DASH-Cro), and MWS (MWS-Cro).

### Instruments

#### Disabilities of the Arm, Shoulder and Hand Questionnaire

The Disabilities of the Arm, Shoulder and Hand (DASH) questionnaire, developed in 1996 by the Upper Extremity Collaborative Group (UECG) (Hudak et al., 1996), was created to address the need for a standardized, region-specific instrument for evaluating upper extremity disability across a wide range of musculoskeletal conditions. The development process involved expert consensus, patient input, and psychometric validation to ensure clinical relevance, reliability, and sensitivity to change across diverse patient populations and treatment modalities. The DASH consists of 30 items grouped into two domains: (1) activity-related difficulty (23 items), which evaluates the extent to which patients experienced difficulty performing specific daily activities during the past week; and (2) symptom severity and social and occupational limitations (7 items), which assess pain, paresthesia, and sleep disturbances, as well as work- and social-functioning restrictions during the same period. Scores range from 0 to 100, with 0 indicating no disability and 100 representing the most severe functional impairment of the upper extremity.

#### Patient-Rated Wrist Evaluation Questionnaire

The Patient-Rated Wrist Evaluation (PRWE) questionnaire is a highly reliable and valid instrument, originally developed in 1996 (MacDermid, 1996). It was designed to address the need for a concise, patient-centered tool specifically targeting wrist pain and functional disability following upper-extremity injuries such as distal radius fractures. The PRWE consists of 15 items divided into two subscales: pain (5 items; maximum score 50, minimum 0) and function (10 items; also scored 0–50). The function subscale is further divided into specific activities (6 items) and usual activities (4 items). Patients rate each item on a 0–10 scale, resulting in a total score ranging from 0 (no pain or disability) to 100 (maximum pain and disability).

#### Mayo Wrist Score

In 1987, Cooney et al. modified the Green and O’Brien score by revising the demerit criteria and eliminating radiographic indices, resulting in what became known as the Mayo Wrist Score (MWS) (Cooney et al., 1987; Dacombe et al., 2016). The MWS is a clinician-administered tool that combines subjective and objective measures to quantify wrist function following trauma or disease. It has been widely used due to its simplicity and its ability to capture both patient-reported symptoms and physical examination findings. The MWS assesses wrist function using four domains: pain intensity (0, 15, 20, or 25 points); functional status - specifically, the ability to return to regular work (0, 15, 20, or 25 points); active range of motion - measuring wrist flexion and extension compared to the contralateral side (0, 5, 10, 15, or 25 points); and grip strength - comparing grip strength of the affected hand to the contralateral side (0, 5, 10, 15, or 25 points). The total possible score ranges from 0 to 100 points and is interpreted as follows: 90–100 points indicate an excellent result; 80–90, a good result; 60–80, a satisfactory result; and below 60, a poor result.

### Statistical analysis

Categorical data are shown by frequency and relative frequency. Quantitative data are presented as mean and standard deviation. Average values of variables that have non-normal distribution (tested with Kolmogorov–Smirnov test) are presented with median and interquartile range.

Before factor anaylsis, we have transformed the values on MWS to z-scores as the items responses have 2 scales (items 1-2 have a 4 point scale and items 3-4 have a 5 point scale). The transformed values were used for the factor analysis and reliability analysis.

We have performed an exploratory factor analysis (EFA), principal axis factoring extraction with oblimin rotation to examine the underlying structure of the measured constructs. Before factor analysis we ran the Kaiser-Meyer-Olkin (KMO) measure of sampling adequacy and the Bartlett’s test of sphericity to test the suitability of the item correlation matrix for factoring. We included the extracted factors with the eigenvalue >1, which accounted for more >10% of the variance, and which passed visual inspection on the scree plot.

Correlations were calculated with Spearman coefficient of correlations as the distributions were not following normal distribution (Shapiro-Wilk test). The strength of correlation was interpreted according to Schober et al (Schober et al., 2018).

P<0.05 value was considered statistically significant. Jamovi version 2.3.28.0. was used for the statistical analysis.

## RESULTS

There were 128 participants and their characteristics are presented in Table 1. Participants were 65 (54-74) years old, mostly female (73%), right handed (52%), dominant hand (54%), treated conservatively (65%).

**Table 1.** Participants’ characteristics.

| Variable | n (%) or C (25-75) |
| --- | --- |
| age | 65 (54-74) |
| Sex |  |
| male | 35 (27) |
| female | 93 (73) |
| Hand |  |
| Left | 55 (43) |
| Right | 66 (52) |
| missing | 7 (5.5) |
| Dominant hand |  |
| Yes | 69 (54) |
| No | 57 (45) |
| missing | 2 (2) |
| Treatment |  |
| Conservative | 79 (62) |
| Operation | 42 (33) |
| Missing | 7 (5.5) |

### Acceptability

The floor and ceiling effects were assessed to evaluate score distribution of the individual items. The DASH items 20 and 21 demonstrated a slight floor effect with more 59% and 61% of the respondents having the lowest score. No ceiling effects were observed.

### Construct validity of DASH, PRWE and MWS

#### Construct validity of DASH

The KMO value (0.94) and the Bartlett’s test of sphericity (PL<L0.001) indicated that factor analysis is appropriate for these data. Principal Axis Factoring (PAF) analysis yielded a two-factor model with 66% of the construct variance explained [eigen values 18.2 (explained variance 46%) and eihgen value 1.3 (explained variance 19%)] (Figure 1). The structure matrix (correlations of each item with the extracted dimensions) and the obtained factorial structure is complex (Table 2). All items including had high loadings (>0.40) indicating strong relationships between the items and the extracted factor but items 17,22 and 23 had factor loadings on both factors. The correlation between the factors is high r=0.73 indicating that this solution is not yielding two factors but 2 facets of the same factor.

**Figure 1.**
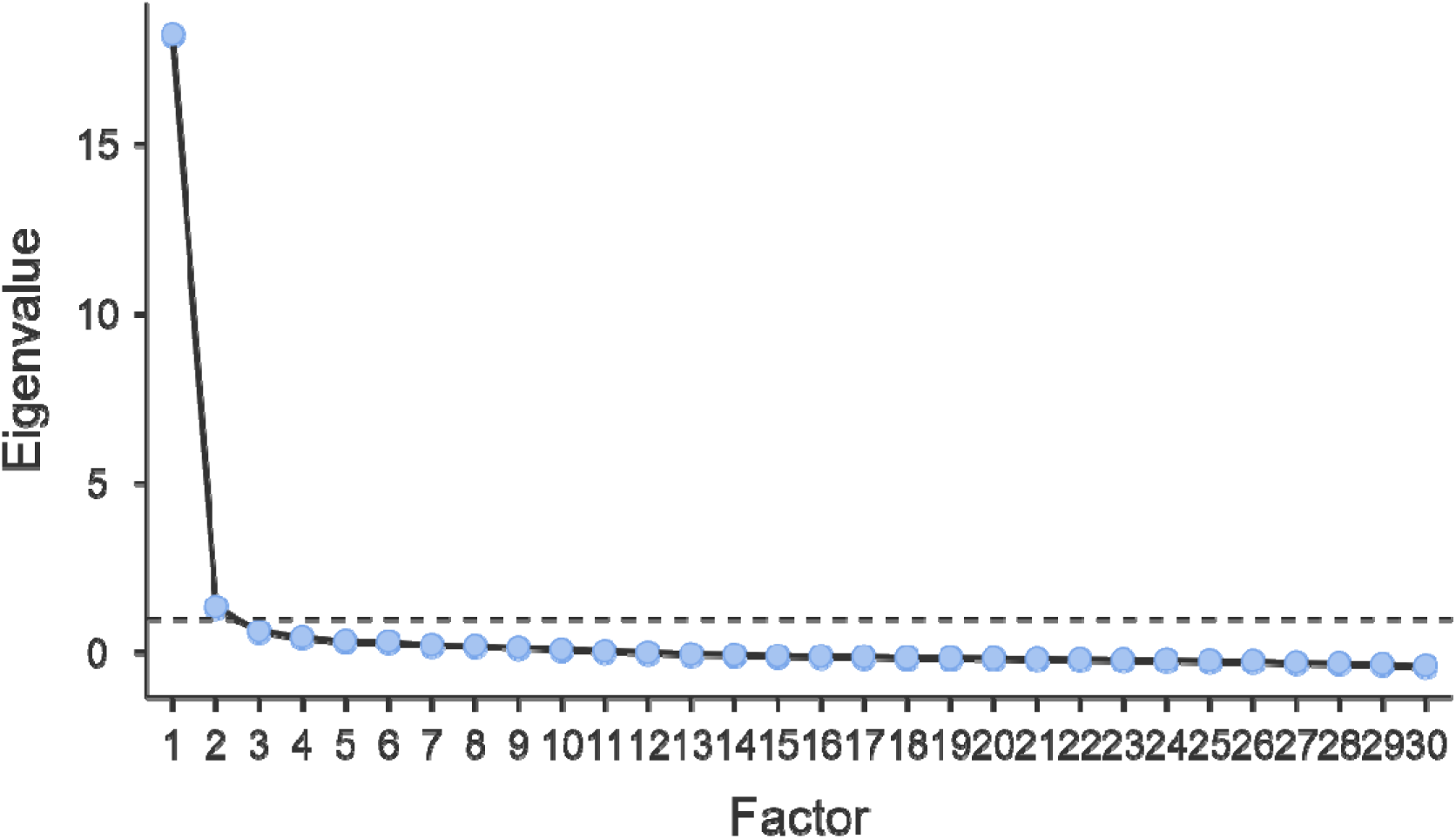
Scree plot of the DASH factor analysis.

**Table 2.**
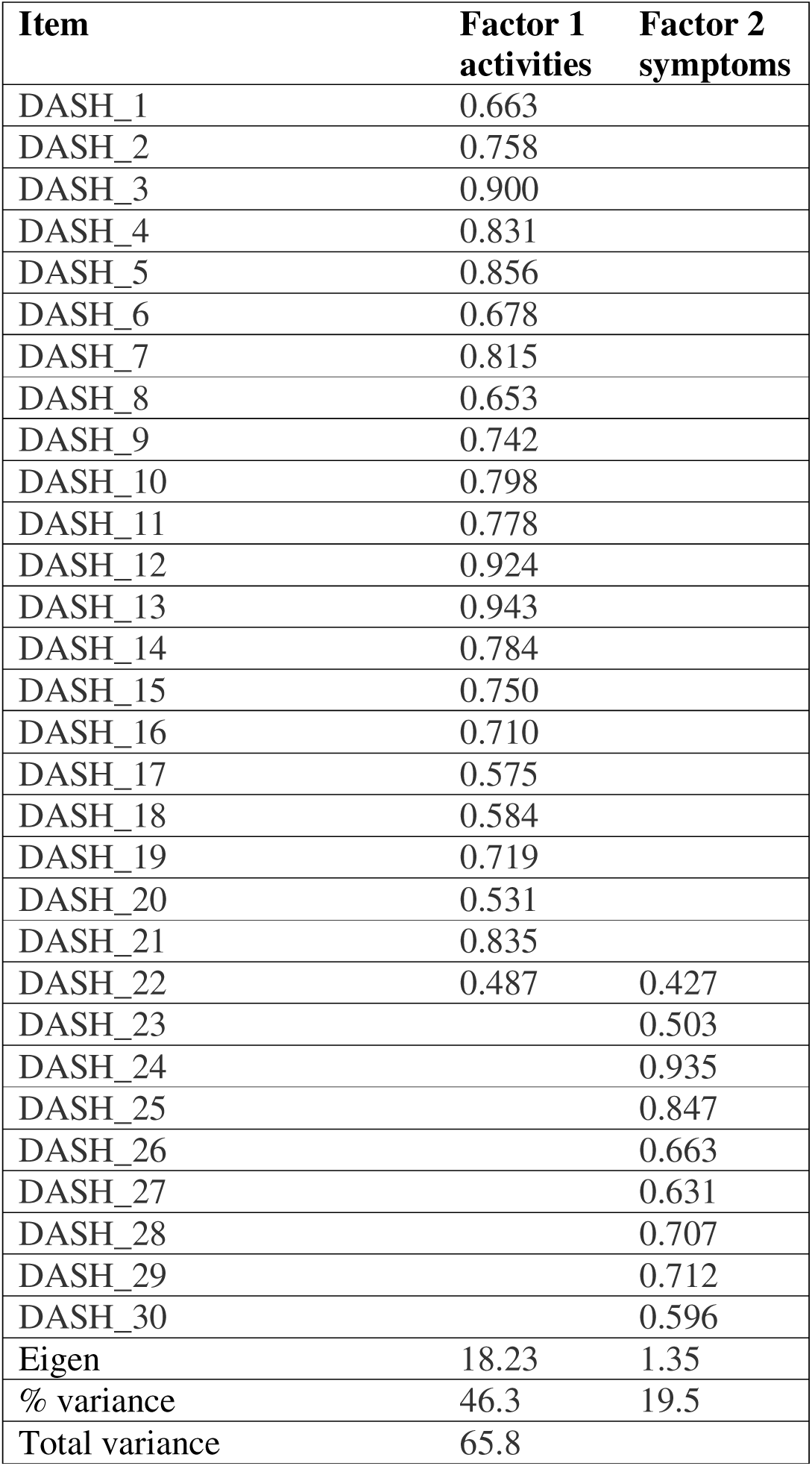
Structure matrices (correlations of each item with the extracted dimensions) of the DASH factor analysis.

#### Construct validity of Quick DASH score

The factor analysis of Quick DASH score is presented in Supplement 1. The KMO value (0.93) and the Bartlett’s test of sphericity (PL<L0.001) indicated that factor analysis is appropriate for these data. Principal Axis Factoring (PAF) analysis yielded a one factor model with 64% of the construct variance explained (eigen value 5.8) (Table S1, Figure 1). The structure matrix (correlations of each item with the extracted dimensions) and the obtained factorial structure is simple. All included items had high loadings (>0.60).

#### Construct validity of PRWE

The KMO value (0.92) and the Bartlett’s test of sphericity (PiJ<iJ0.001) indicated that factor analysis is appropriate for these data. Principal Axis Factoring (PAF) analysis yielded a two factor model (Figure 2) with 75% of the construct variance explained (Table 3). The structure matrix (correlations of each item with the extracted dimensions) and the obtained factorial structure is presented in Table 3 and it corresponds to two factors - function and pain. Those two factors are in high correlation (r=0.71). All items including had high loadings (>0.70) indicating strong relationships between the items and the extracted factors (Table 3).

**Figure 2.**
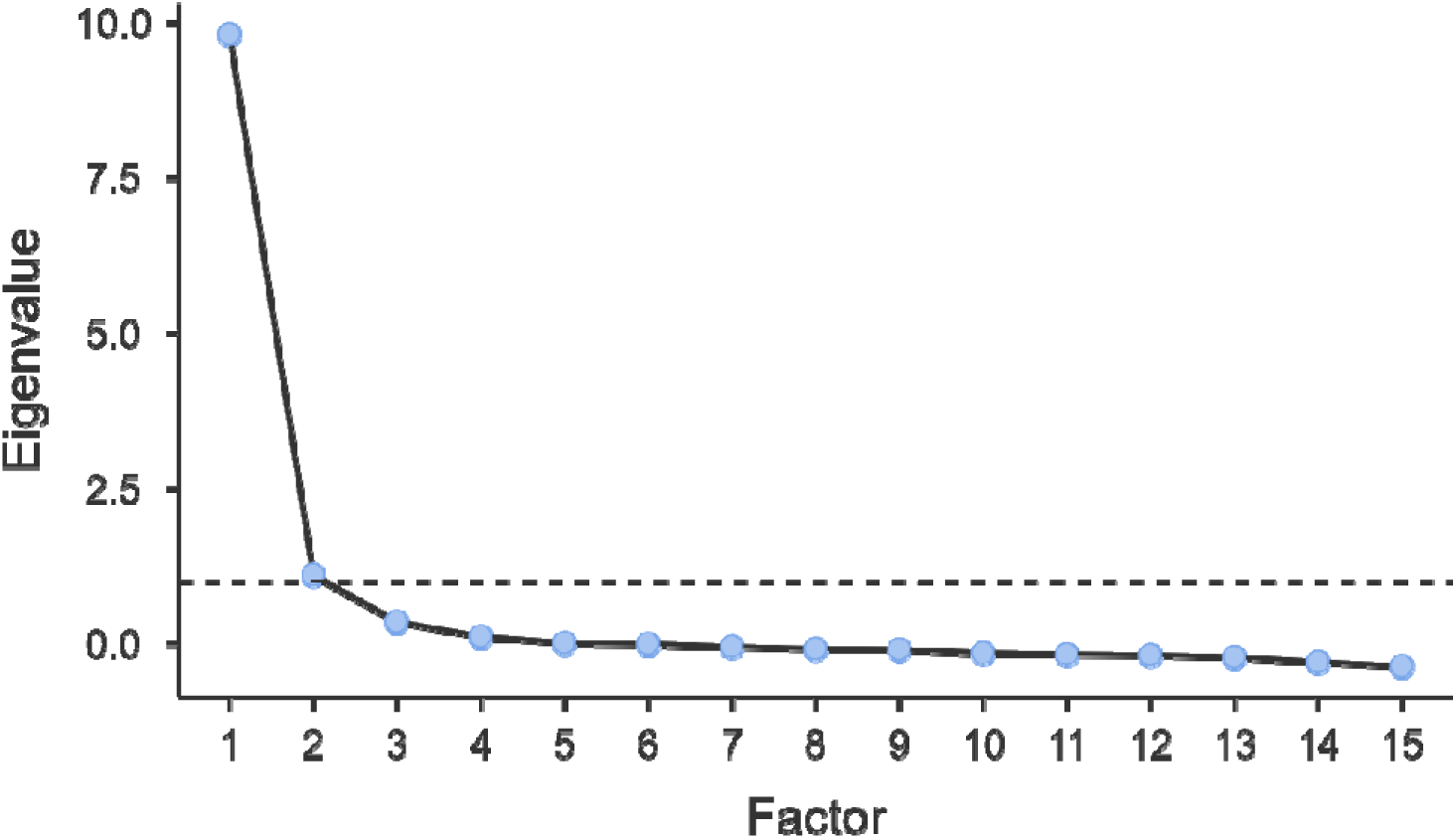
Scree plot of the PRWE factor analysis.

**Table 3.** Structure matrices (correlations of each item with the extracted dimensions) of the PRWE factor analysis.

| Item | Factor 1<br>Function | Factor 2<br>Pain |
| --- | --- | --- |
| PRWE_1 |  | 0.746 |
| PRWE_2 |  | 0.994 |
| PRWE_3 |  | 0.764 |
| PRWE_4 |  | 0.826 |
| PRWE_5 |  | 0.768 |
| PRWE_6 | 0.807 |  |
| PRWE_7 | 0.777 |  |
| PRWE_8 | 0.883 |  |
| PRWE_9 | 0.772 |  |
| PRWE_10 | 0.816 |  |
| PRWE_11 | 0.800 |  |
| PRWE_12 | 0.865 |  |
| PRWE_13 | 0.981 |  |
| PRWE_14 | 0.856 |  |
| PRWE_15 | 0.938 |  |
| **Eigen** | 9.14 | 1.08 |
| **% variance** | 48.9 | 25.8 |
| **Total variance** | 74.7 |  |

Note. 'Principal axis factoring' extraction method was used in combination with a 'oblimin' rotation

#### Construct validity of MWS

The KMO value (0.68) and the Bartlett’s test of sphericity (*P*L<L0.001) indicated that factor analysis is appropriate for the data. Principal Axis Factoring (PAF) analysis yielded a one factor model with 41% of the construct variance explained (eigen value 1.60) (Figure 3). The structure matrix (correlations of each item with the extracted dimensions) and the obtained factorial structure is simple (Table 4). All included items had high loadings (>0.50).

**Figure 3.**
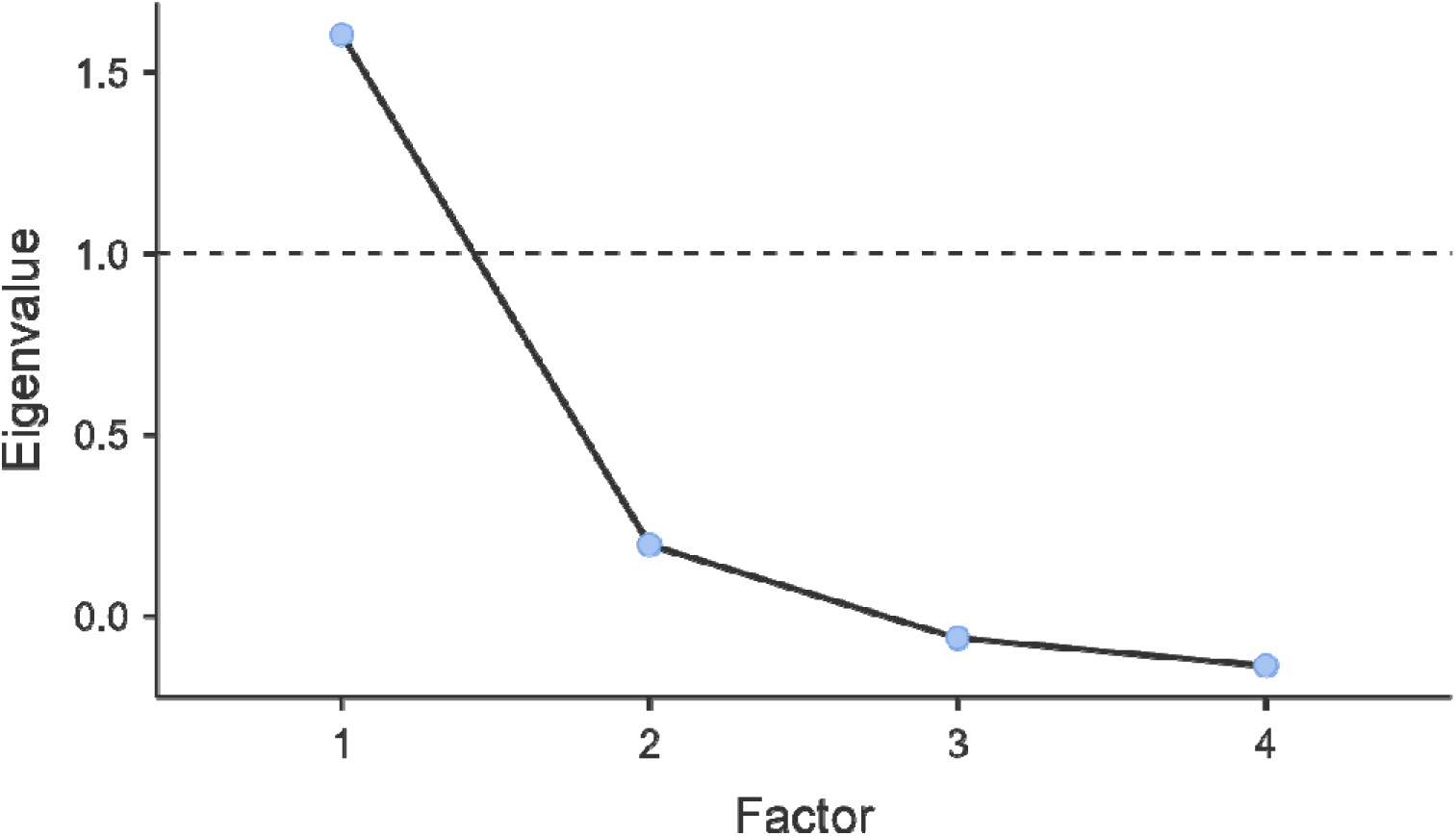
Scree plot of the MWS factor analysis.

**Table 4.**
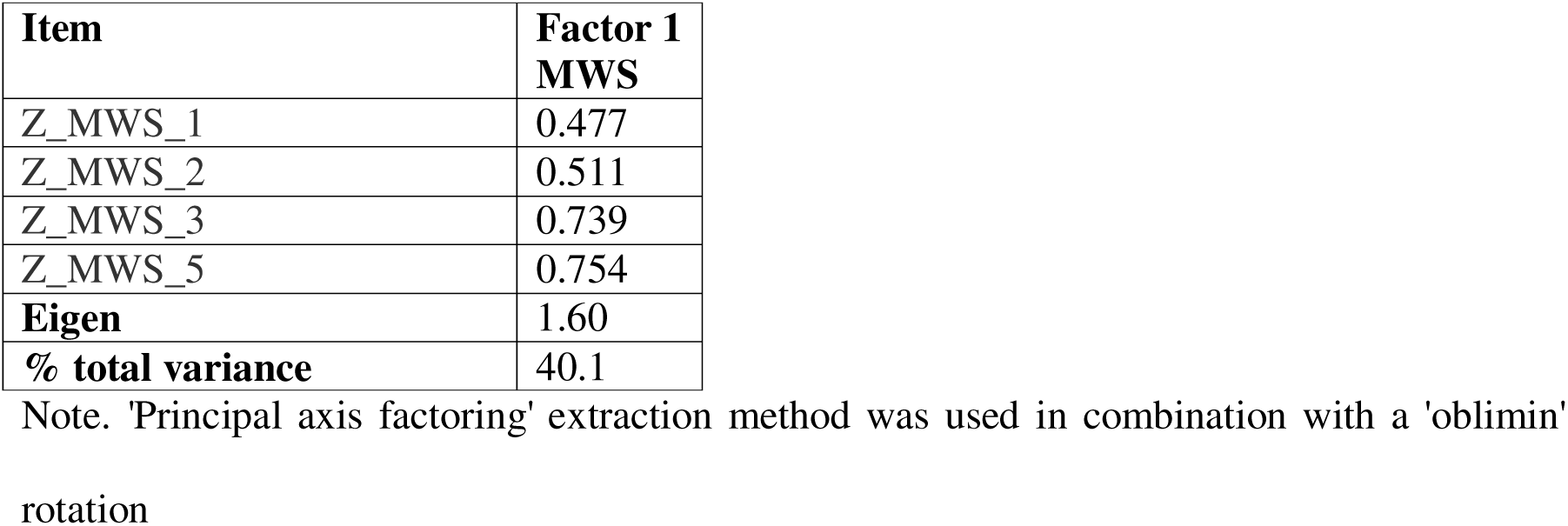
Structure matrices (correlations of each item with the extracted dimensions) of the MWS.

### Reliability analysis of DASH, PRWE and MWS

The overall reliability results are presented in Table 5. The detailed tables are presented in Supplement 1.

**Table 5.** Reliability of the DASH, PRWE and MWS scales and subscales.

| Scale | Reliability<br>Cronbach $\alpha$ |
| --- | --- |
| DASH | 0.98 |
| DASH activities subscale | 0.98 |
| DASH symptoms subscale | 0.92 |
| quick DASH | 0.94 |
| PRWE | 0.97 |
| PRWE function subscale | 0.97 |
| PRWE pain subscale | 0.93 |
| MWS | 0.71 |

#### DASH

The internal consistency of total DASH was very high, Cronbach α=0.98 (Table S2). Both factors have very high reliability, DASH activities subscale – α=0.98 (Table S3) and DASH symptoms – α=0.92 (Table S4). The internal consistency of the quick DASH scale was very high Cronbach α=0.94 (Table S5). The internal consistency of the DASH scales is presented in Table 5 and in detail in supplement 1, Tables S2-S4.

#### PRWE

The internal consistency of the whole PRWE scale was very high Cronbach α=0.97 (Table S6). PRWE function factor had high reliability of 0.97 (Table S7), and PRWE pain had also high reliability of 0.93 (Table S8). Item reliability statistics of PRWE is presented in Table 5 and in detail in supplement 1, tables S6-S8.

#### MWS

The internal consistency of the MWS was acceptable Cronbach α=0.71 (Table S9). Item reliability statistics of MWS is presented in Table 5 and in detail in supplement 1, Table S9.

### Correlations of scales

Correlation of MWS, PRWE, DASH, VAS and age is presented in Table 6. MWS is significantly moderately negatively associated with both PRWE (r_s_=-0.58; p<0.001) and DASH scales (r_s_=-0.61; p<0.001). PRWE and DASH scores are significantly and strongly positively correlated (r_s_=0.71; p<0.001). Age was not in correlation with the MWS, PRWE, DASH scales’ score (all p>0.05). VAS score was significantly negatively weakly correlated with MWS total score (r_s_=-0.36; p<0.001) and positively strongly correlated with PRWE total score (r_s_=0.76; p<0.001) and moderately with DASH total score (r_s_=0.46; p<0.001).

**Table 6.** Correlation of MWS, PRWE, DASH, VAS and age.

|  |  | Age | VAS | MWS_total | PRWE_total | DASH_ukupno |
| --- | --- | --- | --- | --- | --- | --- |
| Age | Spearman's rho | — |  |  |  |  |
|  | df | — |  |  |  |  |
|  | p-value | — |  |  |  |  |
| VAS | Spearman's rho | 0.076 | — |  |  |  |
|  | df | 119 | — |  |  |  |
|  | p-value | 0.407 | — |  |  |  |
| MWS_total | Spearman's rho | -0.128 | -0.364 | — |  |  |
|  | df | 118 | 125 | — |  |  |
|  | p-value | 0.164 | < .001 | — |  |  |
| PRWE_total | Spearman's rho | 0.047 | 0.761 | -0.581 | — |  |
|  | df | 118 | 125 | 124 | — |  |
|  | p-value | 0.608 | < .001 | < .001 | — |  |
| DASH_ukupno | Spearman's rho | 0.006 | 0.457 | -0.606 | 0.708 | — |
|  | df | 119 | 126 | 125 | 125 | — |
|  | p-value | 0.946 | < .001 | < .001 | < .001 | — |

## DISCUSSION

The DASH and PRWE questionnaires are the most commonly used patient-reported outcome measures (PROMs), and numerous studies have confirmed their validity, reliability, and internal consistency for assessing functional outcomes in patients following distal radius fractures (Kleinlugtenbelt et al., 2018). Our study is the first validation of DASH, PRWE and MWS in Croatian language and to the best of our knowledge the first validation of MWS (Wollstein et al., 2025). Both DASH-Cro and PRWE-Cro have excellent construct validtiy and reliability and those results are in line with preivously obtained results of cross-cultural validations (Hemelaers et al., 2008; Öztürk et al., 2015; Alfie et al., 2017; Bastard et al., 2024). MWS-Cro is also a valid instrument, but with lowest validity and reliability probably because of its shortness (4 items) compared to DASH (30 items) and PRWE (15 items).

The DASH questionnaire consists of a total of 30 items divided into two components: an activity component (items 1–23) and a symptom component (items 24–30). Factor analysis of the DASH questionnaire identified two factors that together explained 65.8% of the total variance. Most items were clearly clustered within one of the two factors—functional activity and symptoms—however, minor discrepancies were observed with items 22 and 23. Item 22 (During the past week, to what extent has your arm, shoulder or hand problem interfered with your normal social activities with family, friends, neighbours or groups?) showed moderate factor loadings on both factors (0.487 on the functional factor and 0.427 on the symptom factor), suggesting potential overlap between the functional and symptom components in this item. Item 23 (During the past week, were you limited in your work or other regular daily activities as a result of your arm, shoulder or hand problem?), which theoretically belongs to the functional domain, loaded on the symptom factor, possibly indicating a semantic shift that occurred during the translation process of the questionnaire into Croatian. Apart from these two items, all remaining items demonstrated strong inter-item correlations (ranging from 0.575 to 0.943), as shown in Table 2. Kleinlugtenbelt et al. conducted a similar study in the Netherlands and reported good content validity as well as reliable and internally consistent instruments for evaluating patients with distal radius fractures. Unlike our study, which closely aligned with the original construct, Kleinlugtenbelt et al. identified five factors, explaining less of a total variance (58%), (Kleinlugtenbelt et al., 2018). Vucetic et al. reported results similar to ours, with standardized factor loadings that were statistically significant and ranged from 0.54 to 0.85 (Vucetic et al., 2024). Similarly, de Klerk et al. found high factor loadings for their two-factor model, ranging from 0.597 to 0.896 (de Klerk et al., 2023).

The QuickDASH is a shorter and faster version of the original DASH questionnaire, often considered more practical and user-friendly by clinicians. Results of our factor analysis indicate very good validity (64% variance and one factor structure) and high reliability of the QuickDASH (Cronbach alpha = 0.93). The resulting factorial structure was simple, with all items showing high factor loadings (>0.60), further supporting that the questionnaire measures one dimension related to the functional status of the upper extremity. These findings suggest that the QuickDASH-Cro possesses a stable and unidimensional factor structure and can be considered a suitable instrument for the functional assessment of the arm, shoulder, and hand in clinical setting.

The validated structure of PRWE in our study was consistent with the original, the factors anaylysis yielded 2 factor solution – function and pain. The relaibility of PRWE is very high, for the whole scale (0.97) as well as the subscales (funcion: α=0.97, pain: α=0.93) that are highly correlated indicating in fact unideminesional solution (one factor with 2 facets). PRWE and DASH scores are significantly and strongly positively correlated (r_s_=0.71; p<0.001).

Similarly, Alfie et al. reported a Cronbach’s alpha of 0.96, consistent with findings from other studies (Kim and Kang, 2013; Schønnemann et al., 2013; Alfie et al., 2016). The PRWE (Patient-Rated Wrist Evaluation) was developed as a questionnaire focused solely on the wrist and hand, in contrast to the DASH questionnaire, which assesses the entire upper limb (shoulder, elbow, and hand), and some authors even emphasize its superiority over the DASH, citing that it is faster and easier to administer (MacDermid and Tottenham, 2004; Mellstrand Navarro et al., 2011).

Validaiton of the MWS was done for the first time and with factor analysis of the MWS questionnaire we extracted a single factor, which explained 40.1% of the total variance indicating lower validity. Altough MWS has shown lower validity then DASH and PRWE, we have to take into account that the masure has only 4 items and conclude that this result is satisfying. The reliability of the MWS was also found to be acceptable, with a Cronbach’s alpha of 0.71. MWS was significantly moderately negatively associated with both PRWE (r_s_=-0.58; p<0.001) and DASH scales (r_s_=-0.61; p<0.001). We believe that this results does not preclude MWS useage; on the contrary, from a practical standpoint, MWS can provide a quick and simple screening tool that can later be supplemented by more nuanced instruments like PRWE and DASH.

When selecting the most appropriate questionnaire for assessing the functional status of the hand, it is crucial to balance the psychometric properties of the instrument with its practical applicability. The DASH represents a comprehensive measurement tool that covers the entire upper extremity; however, its relatively lengthy format can be a limitation, especially in situations requiring rapid functional assessment so we advice using the QuickDASH version in clincial setting.

In contrast, the PRWE is specifically focused on the functional evaluation of the hand. Due to its fewer items, ease of administration, and high reliability and validity, the PRWE has proven to be an exceptionally useful tool for routine use, particularly in patients with localized wrist pathologies.

The MWS, being the shortest among the instruments mentioned, allows for a quick and basic assessment of hand functional status. Although less comprehensive, its acceptable reliability makes it valuable in everyday clinical practice when time constraints exist or as a complementary tool alongside more complex questionnaires or diagnostic methods.

## Data Availability

All data produced in the present study are available upon reasonable request to the authors.

## Acknowledgements

None.

## Ethical approval

The study was approved by the medical ethics committees of the University Hospital for Orthopaedics and Traumatology in Lovran, Croatia (reference number 02-249/2021), Faculty of Medicine, University of Rijeka, Croatia (CLASS: 007-08/22-01/25, URNUMBER: 2170-24-04-3/1-22-6), and the and the Kantrida Nursing Home in Rijeka (reference number 01/3972/25).

## Informed consent

Written informed consent was obtained from all participants before the study.

## Supplementary material

Supplemental material for this article is available (supplement 1).

## Author Contributions Statement

BJ, MV, KB and ZJ substantially contributed to: research concept and design, acquisition, analysis, and interpretation of data, drafting the paper and revising it critically, approval of the submitted and final versions. MP substantially contributed to: acquisition of data, drafting the paper and revising it critically, approval of the submitted and final versions. All authors have read and approved the final submitted manuscript.

## REFERENCES

Alfie V, Gallucci G, Boretto J et al. Patient-Rated Wrist Evaluation: Spanish Version and Evaluation of Its Psychometric Properties in Patients with Acute Distal Radius Fracture. J Wrist Surg. 2017, 6: 216–19.

Alfie V, Gallucci G, Boretto J, Donndorff A, De Carli P. Patient Rated Wrist Evaluation: Spanish Version and Evaluation of Its Psychometric Properties. HAND. 2016, 11: 113S–4S.

Aman M, Pennekamp A, Prahm C, Czarnecki P, Van der Heijden B, Harhaus L. The availability of common patient-reported outcome measures in hand surgery across Europe. J Hand Surg Eur Vol. 2025, 50: 681–4.

Bastard C, Sandman E, Balg F, Patenaude N, Chapleau J, Rouleau D. Validity, reliability and responsiveness of the French translation of the Patient-Rated Wrist Evaluation Questionnaire (PRWE). Orthop Traumatol Surg Res. 2024, 110: 103549.

Beaton DE, Wright JG, Katz JN; Upper Extremity Collaborative Group. Development of the QuickDASH: comparison of three item-reduction approaches. J Bone Joint Surg Am. 2005, 87: 1038–46.

Black N. Patient reported outcome measures could help transform healthcare. BMJ. 2013, 346: f167.

Changulani M, Okonkwo U, Keswani T, Kalairajah Y. Outcome evaluation measures for wrist and hand: which one to choose? Int Orthop. 2008, 1: 1–6.

Cooney WP, Bussey R, Dobyns JH, Linscheid RL. Difficult wrist fractures. Perilunate fracture-dislocations of the wrist. Clin Orthop Relat Res. 1987, 214: 136–47.

Court-Brown CM, Caesar B. Epidemiology of adult fractures: A review. Injury. 2006, 37: 691–7.

Dacombe PJ, Amirfeyz R, Davis T. Patient-Reported Outcome Measures for Hand and Wrist Trauma: Is There Sufficient Evidence of Reliability, Validity, and Responsiveness? Hand (N Y). 2016, 11: 11–21.

de Klerk S, Jerosch-Herold C, Buchanan H, van Niekerk L. Structural and cross-cultural validity of the Afrikaans for the Western Cape Disabilities of the Arm, Shoulder and Hand (DASH) questionnaire. J Patient Rep Outcomes. 2023, 7: 1.

Gabel CP, Michener LA, Burkett B, Neller A. The Upper Limb Functional Index: development and determination of reliability, validity, and responsiveness. J Hand Ther. 2006, 19: 328–48.

Hammer OL, Jakobsen RB, Benth JŠ, Randsborg PH. Can Generic Outcome Questionnaires Replace QuickDASH in Monitoring Clinical Outcome Following Surgical Treatment of Distal Radius Fractures? J Hand Surg Am. 2022, 47: 92.e1–92.e9.

Hansen L, Petersen KD, Eriksen SA et al. Subsequent fracture rates in a nationwide population-based cohort study with a 10-year perspective. Osteoporos Int. 2015, 26: 513–19.

Hemelaers L, Angst F, Drerup S, Simmen BR, Wood-Dauphinee S. Reliability and validity of the German version of "the Patient-rated Wrist Evaluation (PRWE)" as an outcome measure of wrist pain and disability in patients with acute distal radius fractures. J Hand Ther. 2008, 21: 366–76.

Hudak PL, Amadio PC, Bombardier C. Development of an upper extremity outcome measure: the DASH (disabilities of the arm, shoulder and hand) [corrected]. The Upper Extremity Collaborative Group (UECG). Am J Ind Med. 1996, 29: 602–8.

Ille M, Matic S, Gambiroza K et al. Assessment of post-traumatic arthritis and functional outcome in patients treated operatively and non-operatively for distal radius Fractures - a 2- year cohort study. Ann Agric Environ Med. 2025, 32: 288–94.

Iskander S, Halbesma G, Hoogbergen MM, Young-Afat D, Veldhuizen IJ. Comprehensive Assessment of Specific Patient-Reported Outcome Measures for Hand and Wrist Conditions in Adults: A Scoping Review. JPRAS Open. 2025, 43: 475–90.

Kim JK, Kang JS. Evaluation of the Korean version of the patient rated wrist evaluation. J Hand Ther. 2013, 26: 238–43.

Kleinlugtenbelt YV, Krol RG, Bhandari M, Goslings JC, Poolman RW, Scholtes VAB. Are the patient-rated wrist evaluation (PRWE) and the disabilities of the arm, shoulder and hand (DASH) questionnaire used in distal radial fractures truly valid and reliable? Bone Joint Res. 2018, 7: 36–45.

Levin LS, Rozell JC, Pulos N. Distal radius fractures in the elderly. JAAOSJournal Am Acad Orthop Surg. 2017, 25: 179–87.

MacDermid JC. Development of a scale for patient rating of wrist pain and disability. J Hand Ther. 1996, 9: 178–83.

MacDermid JC, Khadilkar L, Birmingham TB, Athwal GS. Validity of the QuickDASH in Patients With Shoulder-Related Disorders Undergoing Surgery. J Orthop Sports Phys Ther. 2015, 45: 25–36.

MacDermid JC, Tottenham V. Responsiveness of the disability of the arm, shoulder, and hand (DASH) and patient-rated wrist/hand evaluation (PRWHE) in evaluating change after hand therapy. J Hand Ther. 2004, 1: 18–23

Mellstrand Navarro C, Ponzer S, Törnkvist H, Ahrengart L, Bergström G. Measuring outcome after wrist injury: translation and validation of the Swedish version of the patient-rated wrist evaluation (PRWE-Swe). BMC Musculoskelet Disord. 2011, 12: 171.

Öztürk Ö, Sarı Z, Özgül B, Taşyıkan L. Validity and reliability of the Turkish "Patient-Rated Wrist Evaluation" questionnaire. Acta Orthop Traumatol Turc. 2015, 49: 120–5.

Schober P, Boer C, Schwarte LA. Correlation Coefficients: Appropriate Use and Interpretation. Anesth Analg. 2018, 126: 1763–8.

Schønnemann JO, Hansen TB, Søballe K. Translation and validation of the Danish version of the Patient Rated Wrist Evaluation questionnaire. J Plast Surg Hand Surg. 2013, 47: 489–92.

Tsang P, Walton D, Grewal R, MacDermid J. Validation of the QuickDASH and DASH in Patients With Distal Radius Fractures Through Agreement Analysis. Arch Phys Med Rehabil. 2017, 98: 1217–2.e1.

Tsang S, Royse CF, Terkawi AS. Guidelines for developing, translating, and validating a questionnaire in perioperative and pain medicine. Saudi J Anaesth. 2017, 11: S80–S9.

Vucetic M, Pavlovic V, Milutinovic S et al. Psychometric Properties of the Serbian Version of the Arm, Shoulder, and Hand Disability Self-Assessment Questionnaire: Criterion Validity, Construct Validity, and Internal Consistency. Journal of Clinical Medicine. 2024, 13: 5903.

Wollstein R, Wollstein A, Voegelin E, Ewald S. Challenges in measuring outcomes in the wrist. J Wrist Surg. 2025;eFirst.

