## supplement for "Validation of the Croatian Versions of DASH, PRWE and Mayo Wrist Score in patients with Distal Radius Fractures"

Supplement 1

Factor analysis of DASH quick score

### Exploratory Factor Analysis

|  | | **Factor** |
| --- | --- | --- |
|  | | **1** |
| DASH_1 |  | 0.823 |
| DASH_7 |  | 0.864 |
| DASH_10 |  | 0.794 |
| DASH_14 |  | 0.740 |
| DASH_16 |  | 0.808 |
| DASH_17 |  | 0.802 |
| DASH_22 |  | 0.835 |
| DASH_23 |  | 0.846 |
| DASH_24 |  | 0.687 |

#### Factor Statistics

| Summary | | | | | | |
| --- | --- | --- | --- | --- | --- | --- |
| **Factor** | | **SS Loadings** | | **% of Variance** | | **Cumulative %** |
| 1 |  | 5.78 |  | 64.2 |  | 64.2 |

#### Assumption Checks

| Bartlett's Test of Sphericity | | | | |
| --- | --- | --- | --- | --- |
| **χ²** | | **df** | | **p** |
| 872 |  | 36 |  | < .001 |

| KMO Measure of Sampling Adequacy | | |
| --- | --- | --- |
|  | | **MSA** |
| Overall |  | 0.930 |
| DASH_1 |  | 0.947 |
| DASH_7 |  | 0.920 |
| DASH_10 |  | 0.936 |
| DASH_14 |  | 0.943 |
| DASH_16 |  | 0.938 |
| DASH_17 |  | 0.954 |
| DASH_22 |  | 0.917 |
| DASH_23 |  | 0.902 |
| DASH_24 |  | 0.922 |

#### Eigenvalues

| Initial Eigenvalues | | |
| --- | --- | --- |
| **Factor** | | **Eigenvalue** |
| 1 |  | 5.78237 |
| 2 |  | 0.23172 |
| 3 |  | 0.10325 |
| 4 |  | 0.06006 |
| 5 |  | 0.00762 |
| 6 |  | -0.01068 |
| 7 |  | -0.10438 |
| 8 |  | -0.12089 |
| 9 |  | -0.16629 |

##### Scree Plot

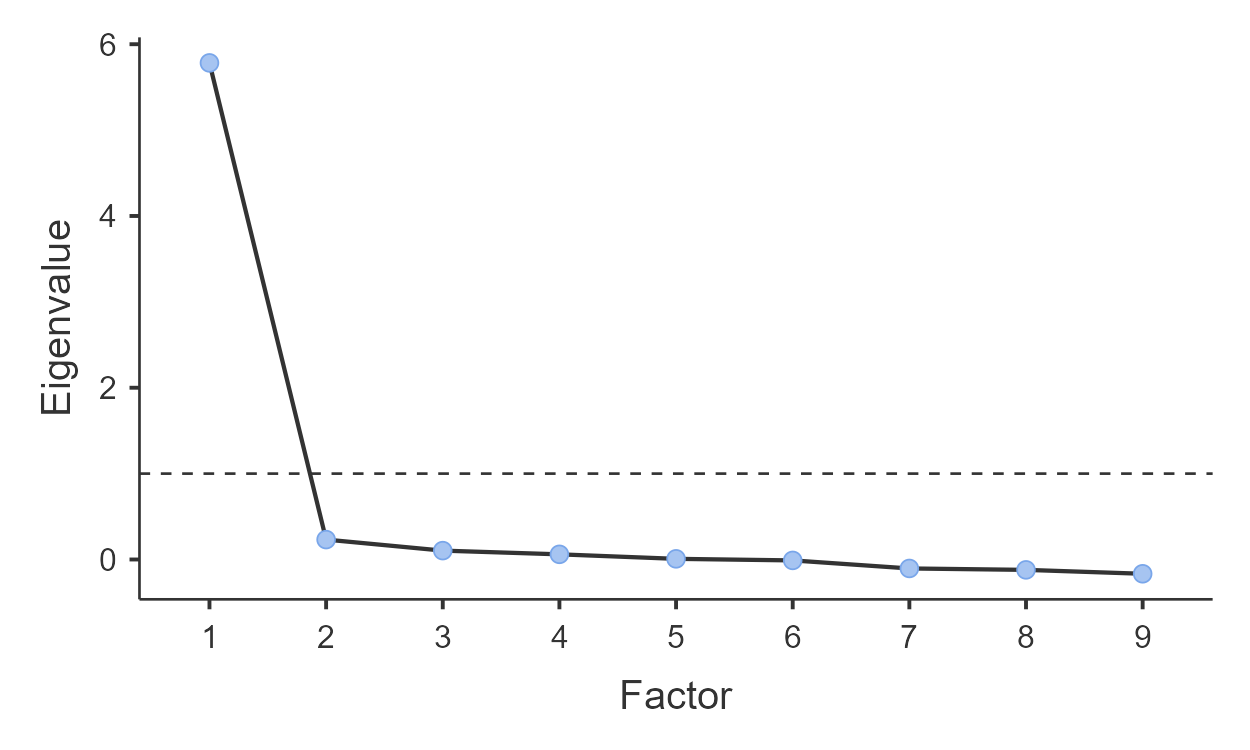

Supplement 2

MWS

Table 1.  MWS item reliability statistics (Cronbach's α=0.71)

| Item Reliability Statistics | | | | | | | | |
| --- | --- | --- | --- | --- | --- | --- | --- | --- |
|  | | | | | | | | **If item dropped** |
|  | | | | **Item-rest correlation** | | | | **Cronbach's α** |
| ZMWS_1 | |  | | 0.423 | | |  | 0.694 |
| ZMWS_2 | |  | | 0.452 | | |  | 0.677 |
| ZMWS_3 | |  | | 0.557 | | |  | 0.613 |
| ZMWS_5 | |  | | 0.567 | | |  | 0.607 |
| Scale Reliability Statistics | | | | | |  |  |  |
|  | | | **Cronbach's α** | | |  |  |  |
| scale |  | | 0.712 | |  |  |  |  |

Table 2. PRWE item reliability statistics (Cronbach's α=0.97)

| Item Reliability Statistics | | | | |
| --- | --- | --- | --- | --- |
|  | | | | **If item dropped** |
|  | | | | **Cronbach's α** |
| PRWE_1 | |  | | 0.966 |
| PRWE_2 | |  | | 0.964 |
| PRWE_3 | |  | | 0.964 |
| PRWE_4 | |  | | 0.965 |
| PRWE_5 | |  | | 0.965 |
| PRWE_6 | |  | | 0.962 |
| PRWE_7 | |  | | 0.962 |
| PRWE_8 | |  | | 0.962 |
| PRWE_9 | |  | | 0.962 |
| PRWE_10 | |  | | 0.963 |
| PRWE_11 | |  | | 0.963 |
| PRWE_12 | |  | | 0.963 |
| PRWE_13 | |  | | 0.962 |
| PRWE_14 | |  | | 0.963 |
| PRWE_15 | |  | | 0.963 |
| Scale Reliability Statistics | | | | |
|  | | | **Cronbach's α** | |
| scale |  | | 0.965 | |

Table 3. PRWE factor 1 - function item reliability statistics (Cronbach's α=0.97)

| Item Reliability Statistics | | | | |
| --- | --- | --- | --- | --- |
|  | | | | **If item dropped** |
|  | | | | **Cronbach's α** |
| PRWE_6 | |  | | 0.963 |
| PRWE_7 | |  | | 0.964 |
| PRWE_8 | |  | | 0.963 |
| PRWE_9 | |  | | 0.964 |
| PRWE_10 | |  | | 0.965 |
| PRWE_11 | |  | | 0.965 |
| PRWE_12 | |  | | 0.965 |
| PRWE_13 | |  | | 0.963 |
| PRWE_14 | |  | | 0.965 |
| PRWE_15 | |  | | 0.964 |
| Scale Reliability Statistics | | | | |
|  | | | **Cronbach's α** | |
| scale |  | | 0.968 | |

Table 4. PRWE factor 2 - pain item reliability statistics (Cronbach's α=0.97)

| Item Reliability Statistics | | | | |
| --- | --- | --- | --- | --- |
|  | | | | **If item dropped** |
|  | | | | **Cronbach's α** |
| PRWE_1 | |  | | 0.926 |
| PRWE_2 | |  | | 0.894 |
| PRWE_3 | |  | | 0.907 |
| PRWE_4 | |  | | 0.907 |
| PRWE_5 | |  | | 0.911 |
| Scale Reliability Statistics | | | | |
|  | | | **Cronbach's α** | |
| scale |  | | 0.926 | |

 Table 5. DASH total item reliability statistics (Cronbach's α=0.97)

| Item Reliability Statistics | | | | |
| --- | --- | --- | --- | --- |
|  | | | | **If item dropped** |
|  | | | | **Cronbach's α** |
| DASH_1 | |  | | 0.977 |
| DASH_2 | |  | | 0.977 |
| DASH_3 | |  | | 0.977 |
| DASH_4 | |  | | 0.977 |
| DASH_5 | |  | | 0.977 |
| DASH_6 | |  | | 0.977 |
| DASH_7 | |  | | 0.977 |
| DASH_8 | |  | | 0.977 |
| DASH_9 | |  | | 0.977 |
| DASH_10 | |  | | 0.977 |
| DASH_11 | |  | | 0.977 |
| DASH_12 | |  | | 0.977 |
| DASH_13 | |  | | 0.977 |
| DASH_14 | |  | | 0.977 |
| DASH_15 | |  | | 0.977 |
| DASH_16 | |  | | 0.977 |
| DASH_17 | |  | | 0.977 |
| DASH_18 | |  | | 0.977 |
| DASH_19 | |  | | 0.977 |
| DASH_20 | |  | | 0.978 |
| DASH_21 | |  | | 0.978 |
| DASH_22 | |  | | 0.977 |
| DASH_23 | |  | | 0.977 |
| DASH_24 | |  | | 0.978 |
| DASH_25 | |  | | 0.977 |
| DASH_26 | |  | | 0.978 |
| DASH_27 | |  | | 0.977 |
| DASH_28 | |  | | 0.977 |
| DASH_29 | |  | | 0.978 |
| DASH_30 | |  | | 0.978 |
| Scale Reliability Statistics | | | | |
|  | | | **Cronbach's α** | |
| scale |  | | 0.978 | |

Table 6. DASH activities item reliability statistics (Cronbach's α=0.98)

| Item Reliability Statistics | | | | |
| --- | --- | --- | --- | --- |
|  | | | | **If item dropped** |
|  | | | | **Cronbach's α** |
| DASH_1 | |  | | 0.974 |
| DASH_2 | |  | | 0.975 |
| DASH_3 | |  | | 0.974 |
| DASH_4 | |  | | 0.974 |
| DASH_5 | |  | | 0.974 |
| DASH_6 | |  | | 0.975 |
| DASH_7 | |  | | 0.974 |
| DASH_8 | |  | | 0.975 |
| DASH_9 | |  | | 0.975 |
| DASH_10 | |  | | 0.974 |
| DASH_11 | |  | | 0.974 |
| DASH_12 | |  | | 0.974 |
| DASH_13 | |  | | 0.975 |
| DASH_14 | |  | | 0.975 |
| DASH_15 | |  | | 0.975 |
| DASH_16 | |  | | 0.974 |
| DASH_17 | |  | | 0.975 |
| DASH_18 | |  | | 0.974 |
| DASH_19 | |  | | 0.975 |
| DASH_20 | |  | | 0.976 |
| DASH_21 | |  | | 0.976 |
| DASH_22 | |  | | 0.975 |
| DASH_23 | |  | | 0.975 |
| Scale Reliability Statistics | | | | |
|  | | | **Cronbach's α** | |
| scale |  | | 0.976 | |

Table 7. DASH symptoms item reliability statistics (Cronbach's α=0.92)

| Item Reliability Statistics | | | | |
| --- | --- | --- | --- | --- |
|  | | | | **If item dropped** |
|  | | | | **Cronbach's α** |
| DASH_24 | |  | | 0.906 |
| DASH_25 | |  | | 0.903 |
| DASH_26 | |  | | 0.914 |
| DASH_27 | |  | | 0.902 |
| DASH_28 | |  | | 0.902 |
| DASH_29 | |  | | 0.910 |
| DASH_30 | |  | | 0.914 |
| Scale Reliability Statistics | | | | |
|  | | | **Cronbach's α** | |
| scale |  | | 0.919 | |

Table 8. Quick DASH factors item reliability statistics (Cronbach's α=0.94)

| Item Reliability Statistics | | | | |
| --- | --- | --- | --- | --- |
|  | | | | **If item dropped** |
|  | | | | **Cronbach's α** |
| DASH_1 | |  | | 0.933 |
| DASH_7 | |  | | 0.932 |
| DASH_10 | |  | | 0.934 |
| DASH_14 | |  | | 0.937 |
| DASH_16 | |  | | 0.934 |
| DASH_17 | |  | | 0.934 |
| DASH_22 | |  | | 0.932 |
| DASH_23 | |  | | 0.932 |
| DASH_24 | |  | | 0.937 |
| DASH_26 | |  | | 0.940 |
| DASH_29 | |  | | 0.938 |
| Scale Reliability Statistics | | | | |
|  | | | **Cronbach's α** | |
| scale |  | | 0.941 | |
